# Evaluation of a novel 5 day accelerated 1 Hz repetitive transcranial magnetic stimulation protocol in major depression: a feasibility study

**DOI:** 10.1101/2020.09.12.20193383

**Authors:** Jean-Philippe Miron, Molly Hyde, Linsay Fox, Jack Sheen, Helena Voetterl, Farrokh Mansouri, Véronique Desbeaumes Jodoin, Ryan Zhou, Sinjin Dees, Arsalan Mir-Moghtadaei, Daniel M. Blumberger, Zafiris J. Daskalakis, Fidel Vila-Rodriguez, Jonathan Downar

**Affiliations:** Krembil Research Institute, University Health Network, Toronto, ON, Canada; Institute of Medical Science, Faculty of Medicine, University of Toronto, Toronto, ON, Canada; Department of Psychiatry, Faculty of Medicine, University of Toronto, Toronto, ON, Canada; Centre Hospitalier de l’Université de Montréal (CHUM) et Centre de Recherche du CHUM (CRCHUM), Département de Psychiatrie, Faculté de Médecine, Université de Montréal, Montréal, QC, Canada; Department of Cognitive Neuroscience, Maastricht University, Maastricht, Limburg, Netherlands; Faculty of Engineering, McMaster University, Hamilton, ON, Canada; Temerty Centre for Therapeutic Brain Intervention at the Centre for Addiction and Mental Health, Toronto, ON, Canada; Department of Psychiatry, University of California San Diego, San Diego, California, USA; Non-Invasive Neurostimulation Therapies Laboratory, Department of Psychiatry, University of British Columbia, Vancouver, BC, Canada

## Abstract

**BACKGROUND:** Repetitive transcranial magnetic stimulation (rTMS) is an effective intervention in major depressive disorder (MDD) but requires daily travel to a treatment clinic over several weeks. Shorter rTMS courses retaining similar effectiveness would thus increase the practicality and scalability of the technique, and therefore its accessibility.

**OBJECTIVE:** We assessed the feasibility of a novel 5 day accelerated 1 Hz rTMS protocol. We hypothesized that this novel rTMS protocol would be safe and well-tolerated while shortening the overall treatment course.

**METHODS:** We conducted a prospective, single-arm, open-label feasibility study. Thirty (30) participants received a one-week (5 days) accelerated (8 sessions per day, 40 sessions total) course of 1 Hz rTMS (600 pulses per session, 50-minute intersession interval) over the right dorsolateral prefrontal cortex (R-DLPFC) using a figure-of-eight coil at 120% of the resting motor threshold (rMT). Primary outcomes were response and remission rates on the Beck Depression Inventory-II (BDI-II).

**RESULTS:** Response and remission rates 1 week after treatment were 33.3% and 13.3% respectively and increased to 43.3% and 30.0% at follow-up 4 weeks after treatment. No serious adverse events occurred. All participants reported manageable pain levels.

**CONCLUSION:** 1 Hz rTMS administered 8 times daily for 5 days is safe and well-tolerated. Validation in a randomized trial will be required.

## INTRODUCTION

Major depressive disorder (MDD) is now the leading cause of disability worldwide, with lifetime suicide rates as high as 15% (Friedrich, 2017; Lam et al., 2016). Even though antidepressant medication offers convenience and simplicity of administration, discontinuation rates are close to 50% at 3 months, resulting from side-effects and lack of clinical response (Kennedy et al., 2016).

Repetitive transcranial magnetic stimulation (rTMS) is well established as an effective intervention in MDD, with an advantageous side-effect profile over medication (Brunoni et al., 2017; Lefaucheur et al., 2020; Milev et al., 2016). Recent meta-analyses report response and remission rates of up to 50-55% and 30-35%, respectively (Milev et al., 2016). Unfortunately, standard rTMS involves treatment courses over several weeks. This complicates treatment logistics for many patients who cannot take time away to attend daily clinic visits for this period of time.

To address this, accelerated rTMS (arTMS), where treatment is delivered multiple times daily, has been studied for over a decade. arTMS has been the subject of several open-label studies (Cole et al., 2020; Dardenne et al., 2018; Fitzgerald et al., 2019a; Holtzheimer et al., 2010; Jodoin et al., 2019; McGirr et al., 2015; Modirrousta et al., 2018; Schulze et al., 2017; Tor et al., 2016; Williams et al., 2018), randomized controlled trials (RCTs) (Baeken, 2018; Baeken et al., 2013; Desmyter et al., 2016; Duprat et al., 2016; Fitzgerald et al., 2018; George et al., 2014; LOO et al., 2007; Theleritis et al., 2017), as well as meta-analyses (Chen et al., 2020; Sonmez et al., 2019). Some evidence suggests that this approach allows comparable effectiveness to standard once-daily rTMS, while shortening treatment length (Fitzgerald et al., 2018). Recently, high-dosage highly-accelerated and personalized intermittent theta-burst (iTBS) arTMS feasibility studies have reported remission rates of up to ∼90%, while delivering treatment over only 5 days (Cole et al., 2020; Williams et al., 2018).

However, arTMS has not been well studied for 1 Hz protocols (Miron et al., 2020a, 2020b). On conventional once-daily regimens, 1 Hz has shown superiority over sham, with some studies also suggesting similar efficacy to HF (Berlim et al., 2012; Brunoni et al., 2017; Lefaucheur et al., 2020; Milev et al., 2016; Miron et al., 2020a). 1 Hz also offers several potential advantages over HF, including less seizure risks (Sun et al., 2012; Vila-Rodriguez et al., 2015), better tolerability (Kaur et al., 2019), and the potential for implementation on simpler, lower-cost equipment (Miron et al., 2020b, 2020a), thus possibly increasing scalability and accessibility.

To address the aforementioned issues, we developed an accelerated low-frequency protocol applying 1 Hz stimulation sessions 8 times daily for 5 days. We hypothesized that the novel protocol would be safe, well-tolerated, and effective, while reducing course length and accelerating clinical improvement.

## METHODS

### Participants

We conducted a prospective, single-center, single-arm, open-label feasibility study. Participants were treated at the Krembil Research Institute, located at the Toronto Western Hospital, an academic healthcare centre which is part of the University Health Network (UHN) in Toronto, Canada. Adult (18-85 years of age) outpatients were included for study participation if they 1) had a Mini International Neuropsychiatric Interview (MINI) confirmed MDD diagnosis (single or recurrent episode) and 2) maintained a stable medication regimen from 4 weeks before treatment start to the end of the study. Exclusion criteria were: 1) history of substance dependence or abuse within the last 3 months; 2) concomitant major unstable medical illness; 3) cardiac pacemaker or implanted medication pump; 4) active suicidal intent; 5) diagnosis of any personality disorder as assessed by a study investigator to be primary and causing greater impairment than MDD; 6) diagnosis of any psychotic disorder; 7) any significant neurological disorder or insult (including, but not limited to: any condition likely to be associated with increased intracranial pressure, space occupying brain lesion, any history of seizure confirmed diagnostically by neurological assessment [except those therapeutically induced by ECT], cerebral aneurysm, Parkinson’s disease, Huntington’s chorea, dementia, stroke, neurologically confirmed diagnosis of traumatic brain injury, or multiple sclerosis); 8) if participating in psychotherapy must have been in stable treatment for at least 3 months prior to entry into the study (with no anticipation of change in the frequency of therapeutic sessions, or the therapeutic focus over the duration of the study); 9) any clinically significant laboratory abnormality in the opinion of the investigator; 10) a dose of more than lorazepam 2 mg daily (or equivalent) currently (or in the last 4 weeks) or any dose of an anticonvulsant due to the potential to limit rTMS efficacy; 11) any non-correctable clinically significant sensory impairment and 12) any significant cardiovascular or metabolic disorder or insult including, but not limited to: coronary artery disease, abnormal heart rhythms, heart failure, cardiac valve disease, congenital heart disease, cardiomyopathy, vascular disease, dyslipidemia, diabetes, or hypertension (this last criteria was added because participants were also enrolled in a cardiac biomarker study, which results will be published elsewhere). All participants provided informed consent and this study was approved by the Research Ethics Board of the University Health Network.

### Study design and procedures

rTMS was delivered through a MagPro R20 stimulator equipped with a MC-B70 coil (MagVenture, Farum, Denmark). Resting motor threshold (rMT) was determined according to standard techniques (McClintock et al., 2017). Treatment consisted of an arTMS course of 8 hourly sessions per day over 5 consecutive weekdays (Monday through Friday), thus totaling 40 sessions in five days. Each rTMS session consisted of low-frequency (LF) 1 Hz stimulation delivered over a 10 min period (1 single train, 600 pulses per session, 50-minute intersession interval) at 120% of rMT over the right dorsolateral prefrontal cortex (R-DLPFC), localized according to a previously published heuristic approximating the F4 EEG site (Mir-Moghtadaei et al., 2017).

Baseline assessments were completed during the week prior to arTMS initiation and consisted of a clinical assessment by trained research staff, including completion of the self-rated Beck Depression Inventory-II (BDI-II) and clinician-rated Hamilton Rating Scale for Depression 17-item (HRSD-17), cap fitting, and motor threshold calibration. Participants were reassessed 1 week and 4 weeks after treatment on the BDI-II and HRSD-17. Participants were asked not to change their medication regimen throughout the whole treatment, up until the 1-week reassessment. Participants who missed any one of the treatment days or 4 or more sessions overall were withdrawn.

To study response trajectory during treatment days, participants also completed the BDI-II at the beginning of each treatment day before rTMS initiation, where they were queried about any adverse events. Participants also completed the BDI-II immediately following their final rTMS session after the last treatment day. Self-rated pain intensity of the rTMS procedure was recorded on a verbal analog scale (VRS – from 0 [no pain] to 10 [intolerable pain]). Moreover, serious adverse events and reasons for treatment discontinuation were recorded when such events occurred. Stimulation intensity was adaptively titrated upward, aiming to reach the target intensity of 120% rMT on the first session of treatment, without exceeding maximum tolerable pain. We recorded the number of sessions required to reach 120% rMT.

### Outcomes

Primary outcome measures were response and remission rates on the BDI-II. Secondary outcomes included score changes and percent improvement. These outcomes were also calculated on the HRSD-17. Response was defined as score reductions of ≥50% from baseline. Remission was defined as a score of ≤12 (Riedel et al., 2010) on the BDI-II and ≤7 on the HRSD-17 (Zimmerman et al., 2004). We also analyzed the outcome trajectories using the BDI-II.

### Statistical analysis

Descriptive statistics were performed on baseline characteristics (age, sex, comorbid anxiety, age of onset of MDD, duration of current MDD episode, total lifetime number of antidepressant medication trials, total ATHF score and baseline BDI score) utilizing independent samples t-tests (two-tailed) for continuous variables, and Chi-square tests for categorical variables. We also performed repeated measures analyses of variance (ANOVA) on BDI-II score at different timepoints to assess the effect of the treatment through time. Planned repeated contrasts were used to make comparisons between the different evaluation times.

## RESULTS

From September 23, 2019 to February 13, 2020, 37 participants with MDD were screened for eligibility, 4 of whom were deemed ineligible or declined to participate; thus, 33 participants were enrolled and began treatment. Of these, 3 discontinued during treatment and were excluded from analysis: 2 participants lost interest and 1 participant was removed by the attending physician after reporting visual symptoms suggestive of possible retinal detachment on day 3 (subsequent diagnosis of migraine equivalent). Thus, 30 participants completed the entire study (**Figure 1**).

**Figure 1:**
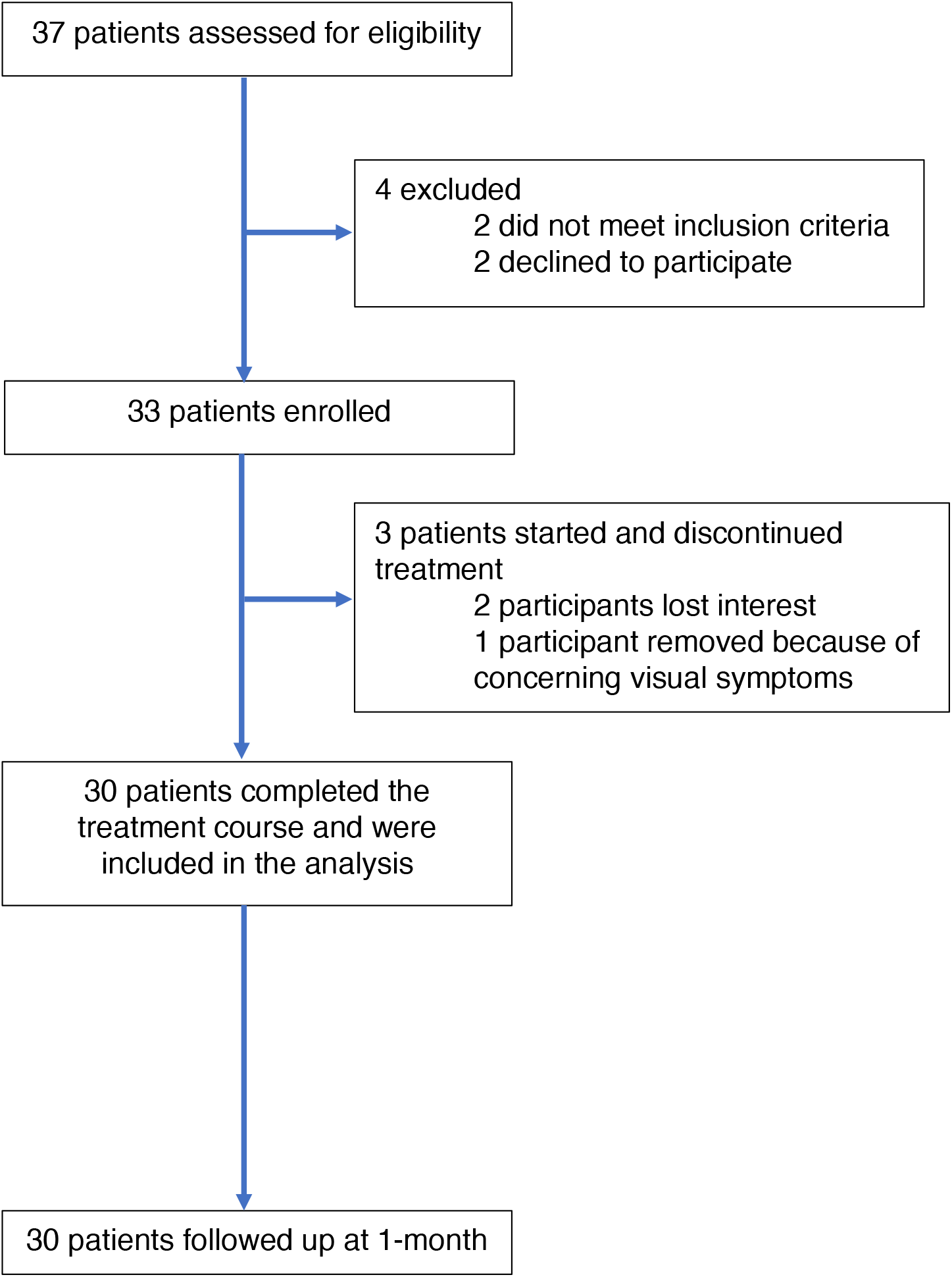
Trial CONSORT flow diagram.

**Table 1** provides the baseline characteristics. Mean age was 43.5 ± 13.9, with 43.3% (13/30) female participants. Mean age of depression onset was 21.4 ± 9.9 years old, with the average length of current episode 13.0 ± 12.7 months. 83.3% of patients were receiving psychopharmacotherapy during the trial, with 60.0% being on at least one antidepressant during the study. Average Antidepressant Treatment History Form (ATHF) total score was 6.6 ± 5.0. The average number of trials on the ATHF in the current episode was 1.3 ± 1.2, with 24/30 (80.0%) having had at least one adequate antidepressant trial in their current depressive episode. Comparing baseline characteristics variables between responders and non-responders did not yield any statistically significant differences (p ≥ .05).

**Table 1:** Baseline characteristics (n = 30).

|  |  |
| --- | --- |
| Age, years | 43.5 (13.9) |
| Women | 43.3% |
| Education, years | 17.1 (3.8) |
| Left-handed | 3.3% |
| Age of onset, years | 21.4 (9.9) |
| Length of current depressive episode, months | 13.0 (12.7) |
| Comorbid anxiety | 70.0% |
| Baseline BDI-II | 35.2 (9.2) |
| Baseline HRSD-17 | 19.8 (4.5) |
| Receiving psychopharmacotherapy during treatment | 83.3% |
| Antidepressant | 60.0% |
| Antidepressant combination | 10.0% |
| Antipsychotic augmentation | 16.7% |
| Psychostimulant augmentation | 30.0% |
| Benzodiazepine | 16.7% |
| ATHF total score | 6.6 (5.0) |
| ATHF number of trials, current episode | 1.3 (1.2) |
| ATHF highest score | 3.3 (1.4) |
| Data are mean (SD) or number of participants (% of total). BDI-II = Beck Depression Inventory-II, HRSD-17 = 17-item Hamilton Rating Scale for Depression, ATHF = Antidepressant Treatment History Form. |  |

Safety and tolerability outcomes are presented in **Table 2**. No serious adverse events (AE) were reported. Overall, 53.3% of patients reported at least one occurrence of an AE at some point during treatment, the most commonly experienced being headache (33.3%). Pain ratings decreased from 3.5 ± 2.0 (first treatment) to 1.7 ± 1.6 (last treatment). Average rMT was 34.6 ± 7.0% of maximum stimulator output, resulting in a mean target stimulation intensity (120%) of 41.6 ± 8.5%. All patients were able to reach their target stimulation intensity, averaging 1.1 ± 0.5 sessions to do so.

**Table 2:**
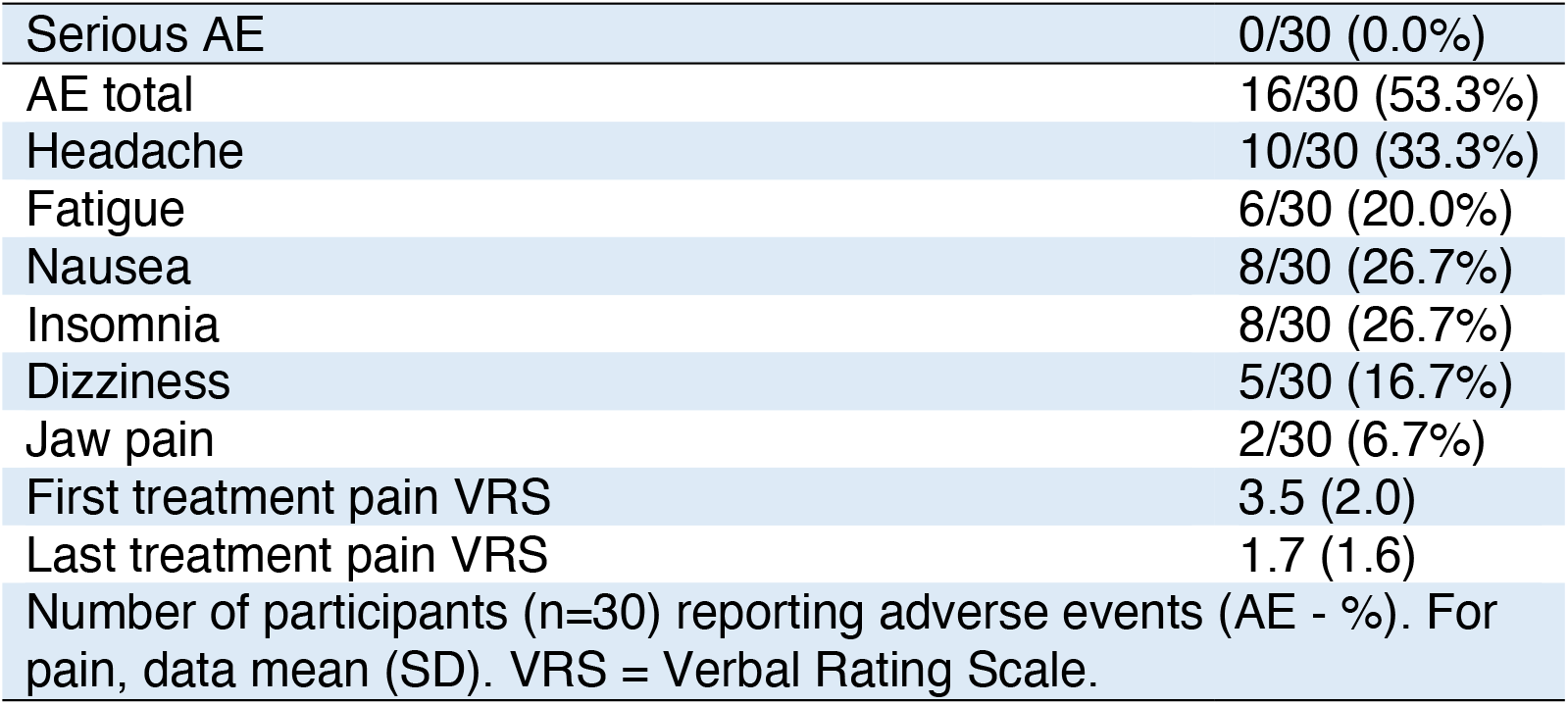
Adverse events.

|  |  |
| --- | --- |
| Serious AE | 0/30 (0.0%) |
| AE total | 16/30 (53.3%) |
| Headache | 10/30 (33.3%) |
| Fatigue | 6/30 (20.0%) |
| Nausea | 8/30 (26.7%) |
| Insomnia | 8/30 (26.7%) |
| Dizziness | 5/30 (16.7%) |
| Jaw pain | 2/30 (6.7%) |
| First treatment pain VRS | 3.5 (2.0) |
| Last treatment pain VRS | 1.7 (1.6) |
| Number of participants (n=30) reporting adverse events (AE - %). For pain, data mean (SD). VRS = Verbal Rating Scale. |  |

**Table 3** presents outcomes of interest. Regarding the primary outcome, response rate was 33.3% (10/30) at 1 week, which increased to 43.3% (13/30) at 4 weeks. Similarly, remission rate increased from 13.3% (4/30) at 1 week to 30.0% (9/30) at 4 weeks. Scores decreased from 35.2 (SD 9.2) at baseline down to 24.0 (11.7) at 1 week, and 23.5 (13.3) at 4 weeks. Percent improvement was 27.5% (32.3%) at 1 week and 33.3% (33.3%) at 4 weeks. Results were similar on the HRSD-17 and are presented in **Table 3**. Also, since we collected daily BDI-II during treatment days, we were able to assess trajectories of outcomes, presented in **Figure 2**. Overall, responders showed rapid improvement during the treatment week, having achieved response on average by the end of the last day, and continued to show slow but steady additional improvement at the 1-and 4-weeks follow-ups (**Figure 2**). Analyses showed a significant improvement in BDI-II score over time (F2,6;54,7 = 13.5, p < 0.001), with planned contrasts showing significant improvements starting on day 2 (p = .018) up to the end of last day of treatment (p = .002), with no further significant changes 1 and 4 weeks after treatment.

**Table 3:**
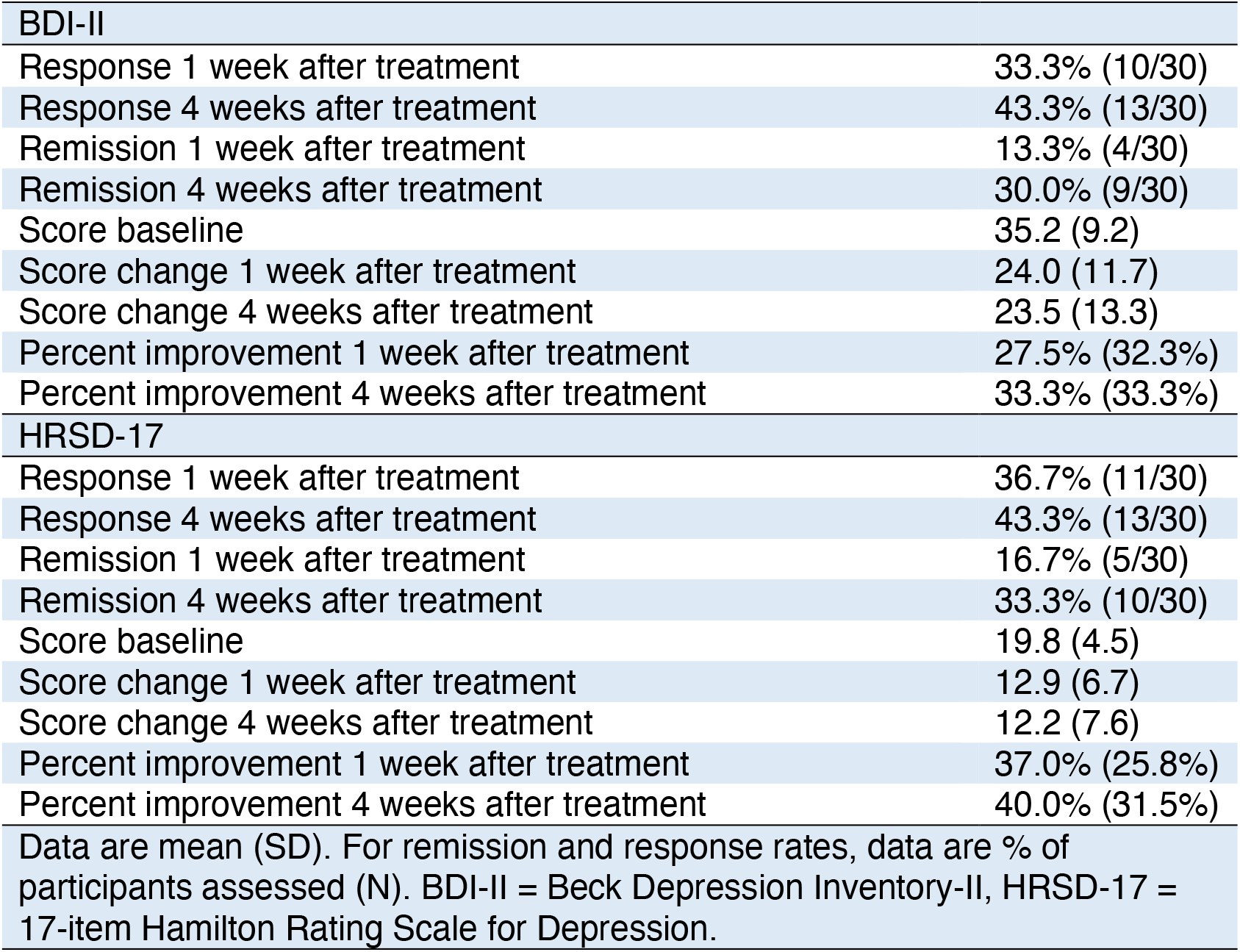
Outcomes of interest

## DISCUSSION

The past three decades have seen the rise of rTMS as an effective and well-tolerated treatment in MDD. Still, conventional once-daily rTMS regimens require frequent visits over 4-6 weeks, thus carrying a travel burden to patients and caregivers. Accelerated protocols, if effective, would reduce travel burden, and offer potential applicability in inpatient or emergency settings.

**Figure 2:**
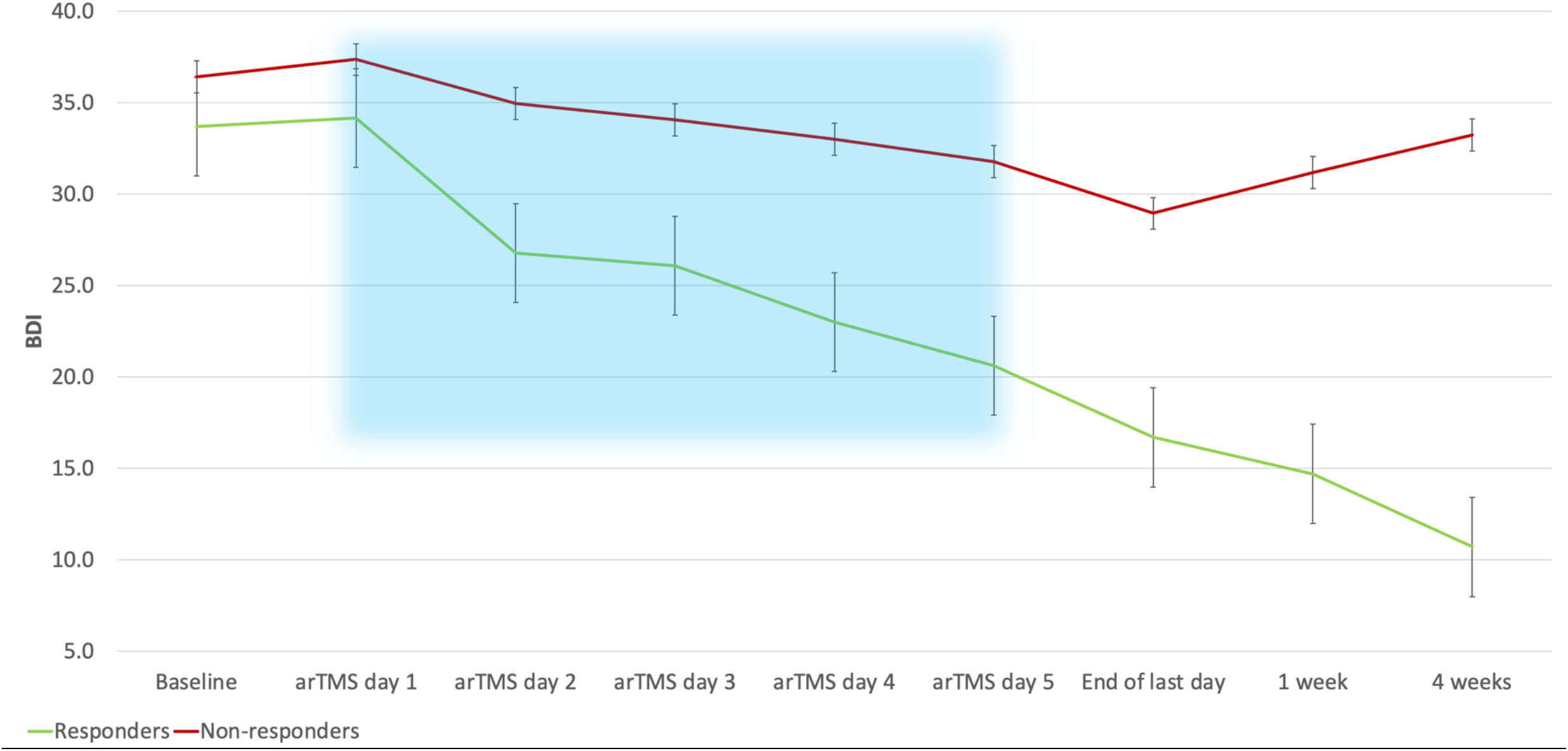
Trajectories of improvement on the BDI-II. Responders showed rapid improvement during the accelerated course, having achieved response on average by the end of the last day, and continued to show slow but steady additional improvement at the 1-and 4-weeks follow-ups. Use of background shading delineates the arTMS course. BDI-II = the Beck Depression Inventory – II, arTMS = accelerated repetitive transcranial magnetic stimulation.

Most accelerated studies to date have employed either high-frequency or intermittent theta-burst stimulation (Baeken et al., 2013; Cole et al., 2020; Fitzgerald et al., 2018; Holtzheimer et al., 2010; LOO et al., 2007). However, 1 Hz right DLPFC protocols have shown better tolerability (Kaur et al., 2019) and similar efficacy to high-frequency left DLPFC protocols in a recent 300-person study on a once-daily regimen (Fitzgerald et al., 2019b), leaving open the question of whether 1 Hz protocols may also be accelerated in a similar fashion. To date, we are only aware of 2 trials having studied 1 Hz arTMS specifically: an initial one was completed in a small patient cohort (N = 7) and used a limited number of sessions (18 over 10 days) (Tor et al., 2016). More recently, our group published another 1 Hz arTMS trial, where 48 participants received 6 daily sessions of 1 Hz arTMS over 5 days (30 sessions total) (Miron et al., 2020b). In this study, which employed a ring-shaped rather than figure-8 coil over F4, we reported modest response and remission rates of 25.0% and 16.7% on the BDI-II 1 week after treatment. Compared to that study, we modified our 1 Hz protocol to increase the number of pulses and daily sessions, in order to potentially maximize treatment effects, switched to a standard figure-8 coil to increase generalizability, and also reassessed at 4 weeks post-treatment without any maintenance or continuation treatment to study if treatment effect could be maintained through time. As in our previous study, response rates at 1 week after treatment were lower than what is usually reported in meta-analyses of standard once-daily rTMS trials (Lefaucheur et al., 2020; Miron et al., 2019), even though the responders subgroup had achieved response on average by the last day of treatment (**Figure 2**). This changed 4 weeks after treatment, where there was a noticeable increase in responders and remitters, reaching 43.3% and 30.0% respectively. This sets our overall number of responders and remitters in the same territory as to what has been reported in large rTMS meta-analyses (Milev et al., 2016). As can be seen in **Figure 2**, a linear trend exists in responders, with improvements seen at every time points. This is supported by our repeated measures ANOVA showing significant improvements already on day 2 of treatment, and maximum response achieved on average by the end of last day of treatment, with a stability at the 1 and 4 weeks follow-ups.

This study has several limitations. Firstly, this was an open-label feasibility study without a sham control arm designed to obtain pilot data for an eventual RCT, where estimates of effectiveness may be more modest. Also, we did not reassess patients between weeks 1 and 4 after treatment, which would have allowed us to establish a more precise trajectory of improvement. In the future, weekly or bi-weekly BDI-II data collection during the follow-up period would be warranted. We also used a limited number of pulses (600 per session), which is 50% lower than what was viewed as maximally efficacious for 1 Hz stimulation in a meta-analysis (Berlim et al., 2012). The rationale behind this was to keep rTMS sessions in the range of ∼10 min, comparable to the 1800-pule iTBS protocol used in the recent Stanford Accelerated Intelligent Neuromodulation Therapy (SAINT) rTMS study (Cole et al., 2020). Of note, 600 pulses are almost twice the amount used in the largest and only multicenter 1 Hz RCT conducted to date (Brunelin et al., 2014). In addition, a recent RCT where high dose 1 Hz rTMS (3600 pulses) was not shown to be more efficacious than standard dose 1 Hz rTMS (1200 pulses) (Fitzgerald et al., 2019b). The optimal number of pulses needed to achieve efficacy with 1 Hz rTMS remains unknown, and future RCTs are required to address this question. In the interim, the practical impediments related to long treatment sessions include the reduced access to rTMS. In this regard, 10 minutes (600 pulses) sessions have clear advantages over 20 minutes (1200 pulses) sessions, allowing 3 to even 4 patients to be treated every hour per machine with the former, compared to 2 patients per hour with the latter. The recent SAINT study also suggested that a high number of daily sessions, spaced by 50 minutes intervals in order maximize long-term potentiation (LTP) mechanisms, might be major parameters in increasing response rates; we thus decided to focus on these aspects in our protocol (Cole et al., 2020). Moreover, we did not require participants to meet the usual requirement of treatment-resistant depression (TRD) in our trial. However, the majority (80%) of participants had failed at least one adequate antidepressant trial in their current depressive episode. There was also no minimum threshold regarding depression severity on the mood scales for study inclusion, but average baseline scores on the BDI-II were in the severe range. Finally, the use of the self-rated BDI-II as our main outcome of interest could be seen as a limitation. However, we also did include outcomes on the HRSD-17 (**Table 3**), which were similar. Self-report scales are a good measure of patient’s perception of their own illness and recovery (Möller, 2000) and the outcomes are unbiased by independent assessors that may skew towards greater improvement in open-label studies. Finally, self-rated scales can be administered daily because of their ease of use, allowing a more fine-grained analysis of outcome trajectories (**Figure 2**) (Möller, 2000).

This feasibility study suggests that a significant proportion of patients may respond rapidly to 1 Hz rTMS, when administered on an accelerated regimen of 8 times daily for 5 days on a standard figure-8 coil. Importantly, we focused on practical considerations in order to facilitate implementation and increase accessibility. Further optimization and validation of the treatment delivery in a formal RCT will be warranted. Finally, such accelerated protocols, shortening treatment courses and thus decreasing the overall number of patients visits to an rTMS clinic, might be a welcomed improvement in our new COVID-19 post-pandemic era.

## Data Availability

Data is available upon reasonable request.

## CONTRIBUTIONS

**JPM** designed the study, was responsible for data collection, analyzed the data, and wrote the manuscript. **MH** and **LF** participated in the study design, data collection, and reviewed the manuscript. **JS** participated in study design, data collection and analysis, and reviewed the manuscript. **HV** participated in study design and reviewed the manuscript. **FM** participated in data analysis and reviewed the manuscript. **RZ** participated in data collection and reviewed the manuscript. **SD, AM, DMB, ZJD** and **FVR** participated in the study design and reviewed the manuscript. **VDJ** participated in data analysis and reviewed the manuscript. **JD** provided resources, supervised all phases of the study and reviewed the manuscript.

## ACKNOWLEDGEMENT

JPM would like to thank the **Brain & Behavior Research Foundation** and the **Branch Out Neurological Foundation** for their financial support of this project. We would like to thank Terri Cairo, Julian Kwok, Meaghan Todd, Nuno Ferreira, Thomas Russell and Eileen Lam for their involvement and organizational support throughout this project. This manuscript has been released as a pre-print at medRxiv (Miron et al., n.d.).

